# SARS-CoV-2 Vaccination Status and MIS-C Incidence: A Systematic Review

**DOI:** 10.64898/2026.05.15.26353349

**Authors:** Katherine Carroll, Hanwen Yang, Ariana Mastrogiannis, Kianna Rojas, Joseph Cervia

## Abstract

Multisystem inflammatory syndrome in children (MIS-C) is a rare but serious condition associated with pediatric SARS-CoV-2 infection. While COVID-19 vaccines prevent infection and reduce severity, less conclusive evidence exists regarding their role in preventing MIS-C during breakthrough infections. This systematic review assessed the impact of SARS-CoV-2 vaccination on MIS-C risk during breakthrough infection. Cross-sectional studies, surveillance studies, and cohort studies were included. Of the 944 studies identified, 6 were included. A significant protective effect was seen in patients who received two doses of SARS-CoV-2 vaccination after exclusion of a biased sample (d= 0.71 [95% CI 0.07 to 1.35; p=0.03]). A trend towards a protective effect was seen after one dose of vaccination, but this effect was not statistically significant. Current literature supports a protective effect of two doses of SARS-CoV-2 vaccination against development of MIS-C in breakthrough COVID-19. The evidence supports clinician advocacy for continued vaccination of children against SARS-CoV-2.

**Article Summary Line:** This systematic review investigates the protective effect of SARSCoV-2 vaccination against development of MIS-C in breakthrough COVID-19 infections.

## Text

Children have not been spared from the impact of the COVID-19 pandemic. One rare but serious complication associated with SARS-CoV-2 infection is the development of multisystem inflammatory syndrome (MIS), or multisystem inflammatory syndrome in children (MIS-C) when occurring in youth, where it is most common. Early in the pandemic, multiple studies documented findings of this condition resembling Kawasaki disease, sharing features of fever, inflammation, and multiorgan system dysfunction *(1-5)*. MIS-C was later recognized as a distinct condition, though the CDC and WHO define it slightly differently. Both definitions include an age limit, which includes patients up to 21 years of age for the CDC and up to 19 years of age for the WHO. Other criteria for both definitions include fever, evidence of multiorgan system damage, elevated markers of inflammation, evidence of exposure to SARS-CoV-2, and the absence of any other plausible diagnoses. MIS-C tends to be severe, with approximately 70% of patients requiring intensive care hospitalization *(6, 7)*.

While the pathophysiology of MIS-C has not yet been fully elucidated, research suggests that the disease may be caused by immune dysregulation as a result of contracting SARS-CoV-2. The disease appears to be a post-infectious process, with cases typically developing 3-6 weeks after exposure to SARS-CoV-2 *(8, 9)*. Many studies have analyzed the serological markers altered during MIS-C. Demonstrated changes include elevations in inflammatory monocyte-activating SARS-CoV-2 IgG antibodies *(10)*, T-cell lymphopenia *(11-13)* and elevated inflammatory cytokines including IFN-γ, IL-1, IL-6, IL-8, and TNF-α *(11, 14, 15)*. Advances in the understanding of the pathogenesis of MIS-C have enabled the development of targeted therapeutics, such as IVIG or IL-1 inhibitors *(15)*. A greater understanding of the underlying pathophysiology of MIS-C is also relevant to the discussion of vaccine effectiveness, such as the possibility of modulating the maturation of CD4 and CD8 T cells and inducing memory T-lymphocyte and B-lymphocyte development *(16)*.

The incidence of MIS-C is not yet fully appreciated, owing in part to the high numbers of asymptomatic COVID-19 cases, particularly in children *(17)*. While multiple vaccines against SARS-CoV-2 have proven effective in preventing COVID-19 and reducing hospitalization rates *(18 - 23)*, few studies have examined whether vaccination lowers the incidence of MIS-C among children who develop breakthrough infections. MIS-C has been shown to be possible to develop even after SARS-CoV-2 vaccination. Previous reviews have not specifically focused on the risk of MIS-C following breakthrough SARS-CoV-2 infection in vaccinated children, nor have they provided a quantitative synthesis of dose-specific effects in this context *(24 - 26)*. The most closely related review by Zhang et al. stratified MIS-C risk by vaccine status, type, and variant, but did not meta-analyze dose-specific protection in breakthrough cases *(24)*. Therefore, this study addresses a critical gap by providing the first systematic review and quantitative synthesis of the protective effect of SARS-CoV-2 vaccination against MIS-C specifically in the context of breakthrough infection.

## Materials and Methods

A systematic search of the electronic databases PubMed, Scopus, and Embase was performed to identify cohort studies reporting the incidence of MIS-C in pediatric patients who had received at least one dose of a SARS-CoV-2 vaccine compared to an unvaccinated patient population. The systematic search included full-text publications from December 2019 through January 2025 using the following search terms: “(vaccine* OR “Vaccination”[Mesh]) AND (MISC OR “MIS-C” OR “multisystem inflammatory syndrome in children” OR “multisystem inflammatory syndrome” OR “pediatric multisystem inflammatory disease, COVID-19 related” [Supplementary Concept])”. The papers of interest also had to include a reference cohort of SARS-CoV-2 positive, MIS-C negative patients as a control group. The reference lists of identified studies were examined to identify further reports of interest.

It is important to note that inclusion criteria was limited to studies that provided evidence of MIS-C after breakthrough SARS-CoV-2 infection, either through positive nucleocapsid serology or a positive PCR confirming SARS-CoV-2 infection. Studies that focused on a hyperinflammatory response to SARS-CoV-2 vaccination were excluded. Similarly, studies that began by using a screening program to identify all MIS-C cases often could not be included, as they lacked a reference cohort of SARS-CoV-2 positive patients. When possible, authors were contacted for further information, and the studies were included if data was subsequently made available *(27 - 31)*.

### Inclusion and exclusion criteria

Inclusion criteria were: (1) study type: cohort and surveillance studies (2) participants: children in the health care and/or community settings with a diagnosis of COVID-19 or MIS-C; (3) intervention and comparator: SARS-CoV-2 immunized vs non-immunized patients; (4) primary outcome: MIS-C diagnosis following the CDC guideline; (5) settings: hospital or community.

Exclusion criteria were: (1) theoretical models; (2) nonhuman experimental laboratory studies; (3) conference abstracts; (4) non-English language publication (5) studies lacking in evidence of breakthrough SARS-CoV-2 infection through positive nucleocapsid serology or positive PCR test.

### Study selection and data extraction

Two reviewers (KC and HY) independently screened the articles based on the titles, abstracts, and full texts. Then, two reviewers (KC and HY) independently extracted the following data from included studies: first author, publication year, country, disease, details of study population and intervention, study design, sample size, settings, and results. All disagreements were resolved by discussion and consensus. Articles that were considered included cross-sectional studies, surveillance studies, and cohort studies. Only peer-reviewed or pre-print studies were considered. These data were analyzed to discern associations between SARS-CoV-2 immunization of any type and the severity of clinically diagnosed MIS-C. The initial review yielded 415 studies. Title and abstract screening, as well as full-text review of each manuscript, were conducted by each of the two reviewers (KC and HY), and any disagreement was resolved by discussion and consensus.

All search results were imported into Covidence Systematic Review Software (Veritas Health Innovation Ltd, Melbourne VIC 3000, Australia) for review. The literature was reviewed based on pre-established inclusion and exclusion criteria described above by two independent reviewers. The literature search was rerun, using the same search terms and methods mentioned above, twice to ensure most recently published journals were included in the review. The first search was completed on July 22^nd^ 2022 and the second search on January 1^st^ 2023. A third search was completed on July 3^rd^ 2025.

Sixty-five studies were found to meet the inclusion criteria, and after full-text review, 6 studies that reported SARS-CoV-2 immunization status and outcome data were used in the final analysis (Figure 1) *(30 - 35)*. Frequencies, percentages, relative risk, confidence intervals, and t-tests were calculated as appropriate, to measure differences between groups. Additionally, all studies included were summarized in abbreviated form and categorized by date of publication, country, site, proportion of infected subjects, and main findings (Table 1). All data were compiled and analyzed using Microsoft Excel.

**Table 1:** Study characteristics included in final meta-analysis.

| Study, year, site | Study design | Start Date – End Date | Sample Size | Percent Vaccinated (%) | Youngest Vaccine Eligible at Study Start, Youngest Eligible at Study End | Vaccine Definition | Median Age | Main Findings |
| --- | --- | --- | --- | --- | --- | --- | --- | --- |
| Kildegaard (2022) Denmark | Cohort Study | Feb 20 – Oct 21 | 70666 | 2.65 | n/a, 12-15y | 2 doses | 8 | Vaccination with two doses was associated with a significantly lower risk of MIS-C compared to unvaccinated children. |
| LaRovere (2023) US | Surveillance | Dec 20 – Dec 21 | 2168 | 1.25 | 16-17y, 5-11y | Partial (1) and Full (2) | 10.3 | Incidence of MIS-C was markedly lower in partially and fully vaccinated children; most cases occurred among unvaccinated patients. |
| Bahl (2023) US | Cohort Study | Jan 21– Jun 22 | 566 | 10.25 | 16-17y, 6 mo- 5y | Partial (1), Full (2), and boosted | 5.2 | Vaccination (including partial, full, and boosted) was associated with a reduced risk of MIS-C hospitalization; no MIS-C cases occurred in boosted children. |
| Wanga (2021) US | Surveillance | Jul 21– Aug 21 | 915 | 2.3 | 12-15y, 12-15y | Partial (1) and Full (2) | 8 | Very low proportion of vaccinated MIS-C cases; small number of vaccinated children limited statistical power to assess risk reduction. |
| Nygaard (2022) Denmark | Cohort Study | Aug 21– Feb 22 | 92569 | 7.1 | 12-15y, 5-11y | 2 doses | 8 | Two-dose vaccination significantly reduced the risk of MIS-C compared to no vaccination. |
| Schwartz (2024) Israel | Cohort Study | Apr 20 – Nov 21 | 526685 | 2.7 | 16y, 12y | 2 doses | *12 | Vaccination with two doses was associated with a markedly decreased incidence of MIS-C in SARS-CoV-2 positive children. |

\*The median age was not reported for this study; 12 years is the mean age

| Study, year, site | Study design | Start Date - End Date | Sample Size | Percent Vaccinated (%) | Youngest Vaccine Eligible at Study Start, Youngest Eligible at Study End | Vaccine Definition | Median Age | Main Findings |
| --- | --- | --- | --- | --- | --- | --- | --- | --- |
| Kildegaard (2022) Denmark | Cohort Study | Feb 20 – Oct 21 | 70666 | 2.65 | n/a, 12-15y | 2 doses | 8 | Vaccination with two doses was associated with a significantly lower risk of MIS-C compared to unvaccinated children. |
| LaRovere (2023) US | Surveillance | Dec 20 – Dec 21 | 2168 | 1.25 | 16-17y, 5-11y | Partial (1) and Full (2) | 10.3 | Incidence of MIS-C was markedly lower in partially and fully vaccinated children; most cases occurred among unvaccinated patients. |
| Bahl (2023) US | Cohort Study | Jan 21- Jun 22 | 566 | 10.25 | 16-17y, 6 mo- 5y | Partial (1), Full (2), and boosted | 5.2 | Vaccination (including partial, full, and boosted) was associated with a reduced risk of MIS-C hospitalization; no MIS-C cases occurred in boosted children. |
| Wanga (2021) US | Surveillance | Jul 21- Aug 21 | 915 | 2.3 | 12-15y, 12-15y | Partial (1) and Full (2) | 8 | Very low proportion of vaccinated MIS-C cases; small number of vaccinated children limited statistical power to assess risk reduction. |
| Nygaard (2022) Denmark | Cohort Study | Aug 21- Feb 22 | 92569 | 7.1 | 12-15y, 5-11y | 2 doses | 8 | Two-dose vaccination significantly reduced the risk of MIS-C compared to no vaccination. |
| Schwartz (2024) Israel | Cohort Study | Apr 20 – Nov 21 | 526685 | 2.7 | 16y, 12y | 2 doses | *12 | Vaccination with two doses was associated with a markedly decreased incidence of MIS-C in SARS-CoV-2 positive children. |

\*The median age was not reported for this study; 12 years is the mean age

**Figure 1:**
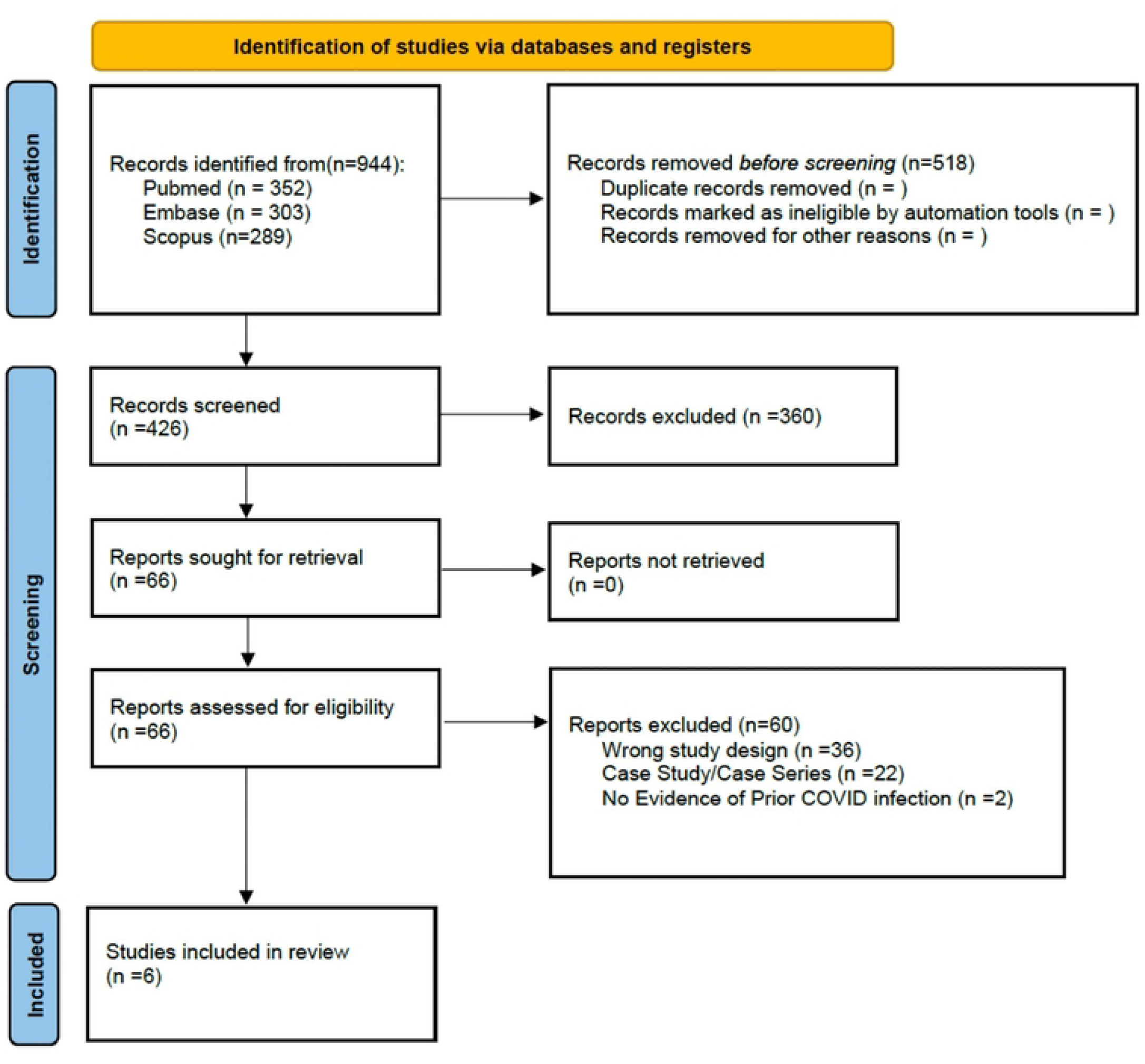
PRISMA flow chart of systemic review.

Of note, there are a few papers that merit explanations for their exclusion from our analysis. The paper “BNT162b2 mRNA Vaccination Against Coronavirus Disease 2019 is Associated With a Decreased Likelihood of Multisystem Inflammatory Syndrome in Children Aged 5–18 Years—United States, July 2021 – April 2022” by Zambrano et al. *(36)* used a SARS-CoV-2 negative control group, rather than a SARS-CoV-2 positive control group, which was needed for analysis of breakthrough MIS-C cases in COVID-19 positive individuals. The paper by Levy et al. *(27)* also did not include a SARS-CoV-2 positive control group. Finally, the paper “Risk and Phenotype of Multisystem Inflammatory Syndrome in Vaccinated and Unvaccinated Danish Children Before and During the Omicron Wave” by Dr. Holm used the same MIS-C cases as reported in the paper by Dr. Kildegaard (confirmed by Dr. Kildegaard) and consequently could not be included without skewing the analysis by counting these cases twice.

### Data Analysis

The primary outcome variable of interest was the log odds ratio of vaccination effect on MIS-C incidence, and the overall effect size was weighted by the sample size of each study. The confidence interval was obtained using 95% confidence. Sample selection bias was analyzed using funnel plot as well as heterogeneity analysis.

Data were separated into 3 subsets, tabulating data from at least one dose of SARS-CoV-2 vaccination, at least two doses of vaccination, and data from the unvaccinated cohort. Analysis was performed twice, once comparing patients having received at least one dose of a SARS-CoV-2 vaccine to unvaccinated cohorts and once comparing the cohort with two or more vaccinations to patients receiving one or no doses of a vaccine. Meta-analysis was performed using IBM SPSS Statistics (Version: 29.0.0.0 (241)). Graphs were generated using IBM SPSS statistics.

### Quality Appraisal and Risk of Bias

Risk of bias analysis was performed using Mixed methods appraisal tool (MMAT) Version 2018. Appraisal was conducted by two reviewers

## Results

### Study characteristics

Of the 6 studies included in the meta-analysis, 3 studies were from the United States, 2 were from Denmark, and 1 was from Israel. Four of the studies included were cohort studies while 2 were surveillance studies.

### Bias analysis

Funnel plot analysis is designed to check for publication bias, plotting effect size against study precision. This analysis assumes that a more precise study will fall closer to the average result, and therefore a “well-behaved” study selection without bias will fall under an inverted funnel region. Heterogeneity analysis is a calculation of variability between studies, which in a meta-analysis consisting of similar study protocol and outcome, should only be due to measurement error alone. Thereby, a non-zero heterogeneous effect represents uncertainty in the overall result.

### One-Dose of Vaccine Odds Ratio

Three studies reported the effect of at least one dose of the COVID-19 vaccine compared to no doses of the vaccine. The average effect size was not significant, d = 0.39 [95% CI -0.17 to 0.95; p=0.17]. The heterogeneity test showed I^2^ = 0%, indicating no detectable heterogeneity of the results. There was no evidence of systemic publication bias on inspection of a funnel plot. (Figure 2).

**Figure 2a.**
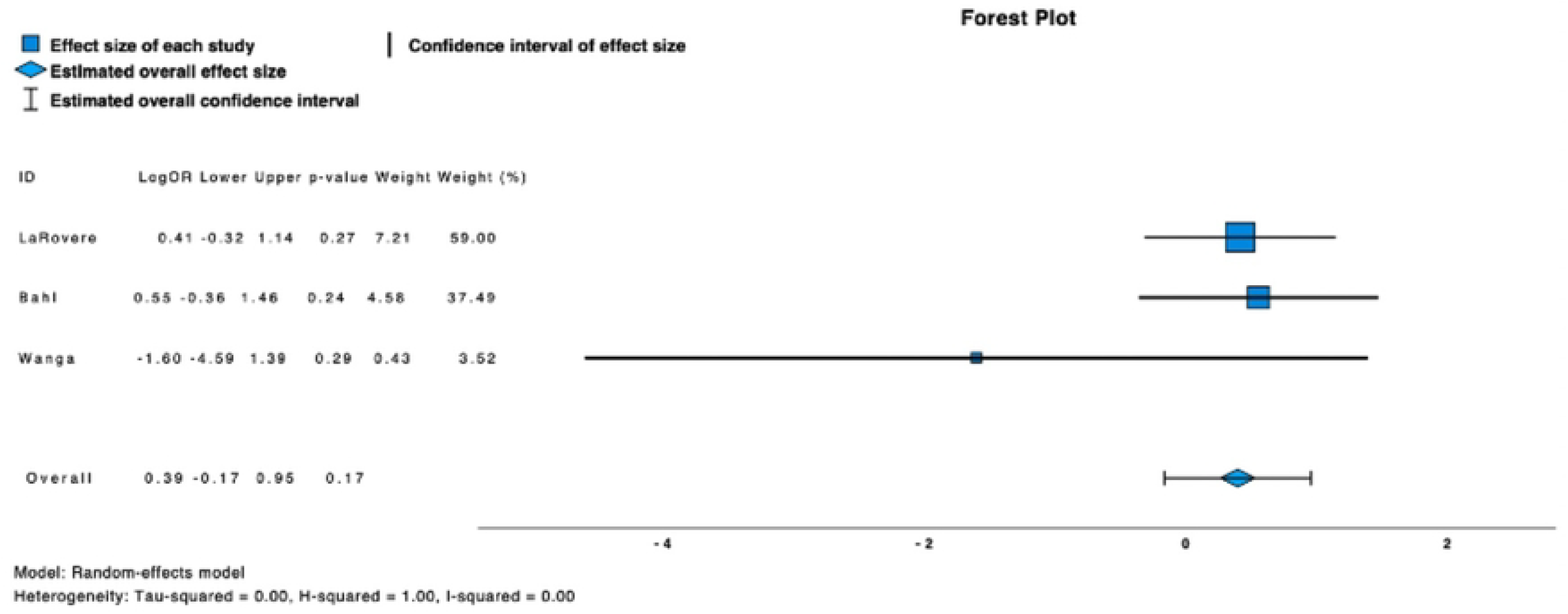
Forest plot of the effect of one vaccination on development of MIS-C, showcasing an effect size of 0.39 (-0.17∼0.95).

**Figure 2b.**
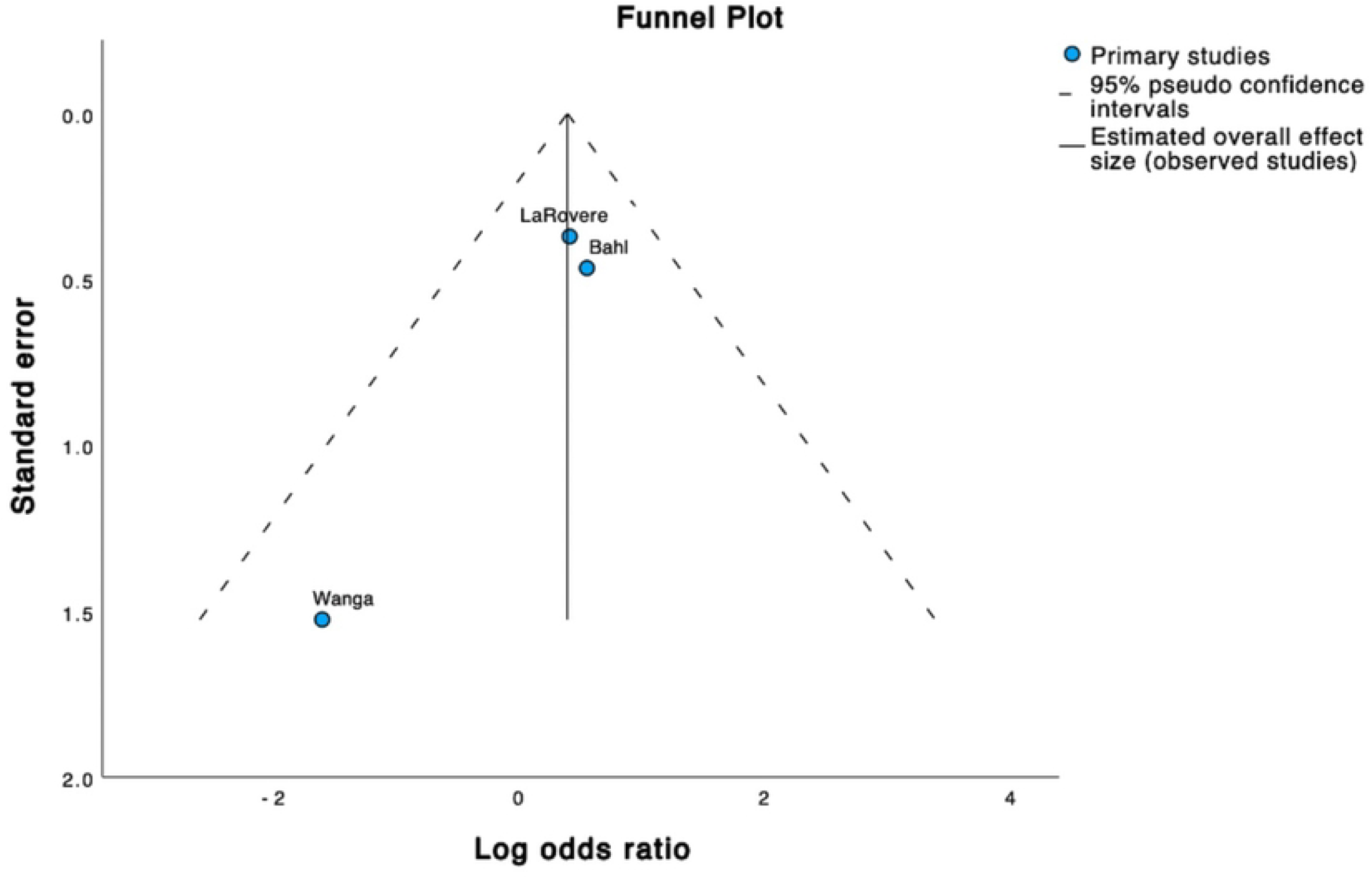
Funnel plot of the effect of one vaccination on development of MIS-C, displaying a symmetric graph implying low publication bias.

### Two Doses of Vaccine Odds Ratio

Six studies reported the effect of at least two doses of the COVID-19 vaccine compared to either one or zero doses of the vaccine. The average effect size was not significant, d = 0.36 [95% CI -1.09 to .357; p=0.32]. Funnel plot analysis showed a tau-squared of 0.23 and I^2^ = 28%, indicating a moderate heterogeneity effect. A funnel plot suggested a potential outlier in the Wanga et al. study (Figure 3).

**Figure 3a.**
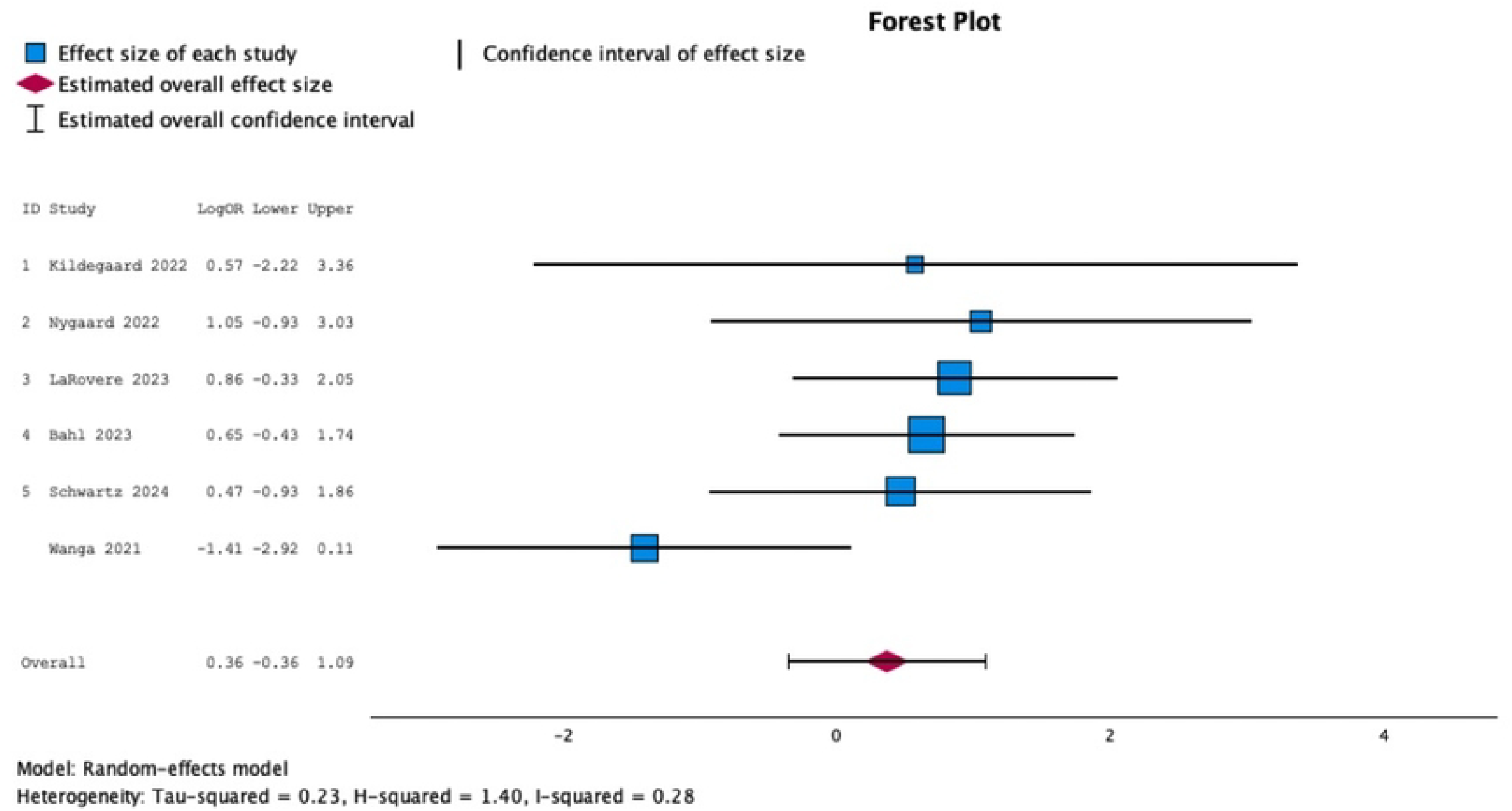
Forest plot of the effect of two vaccinations on development of MIS-C, showcasing an effect size of 0.34 (-0.58∼1.25).

**Figure 3b.**
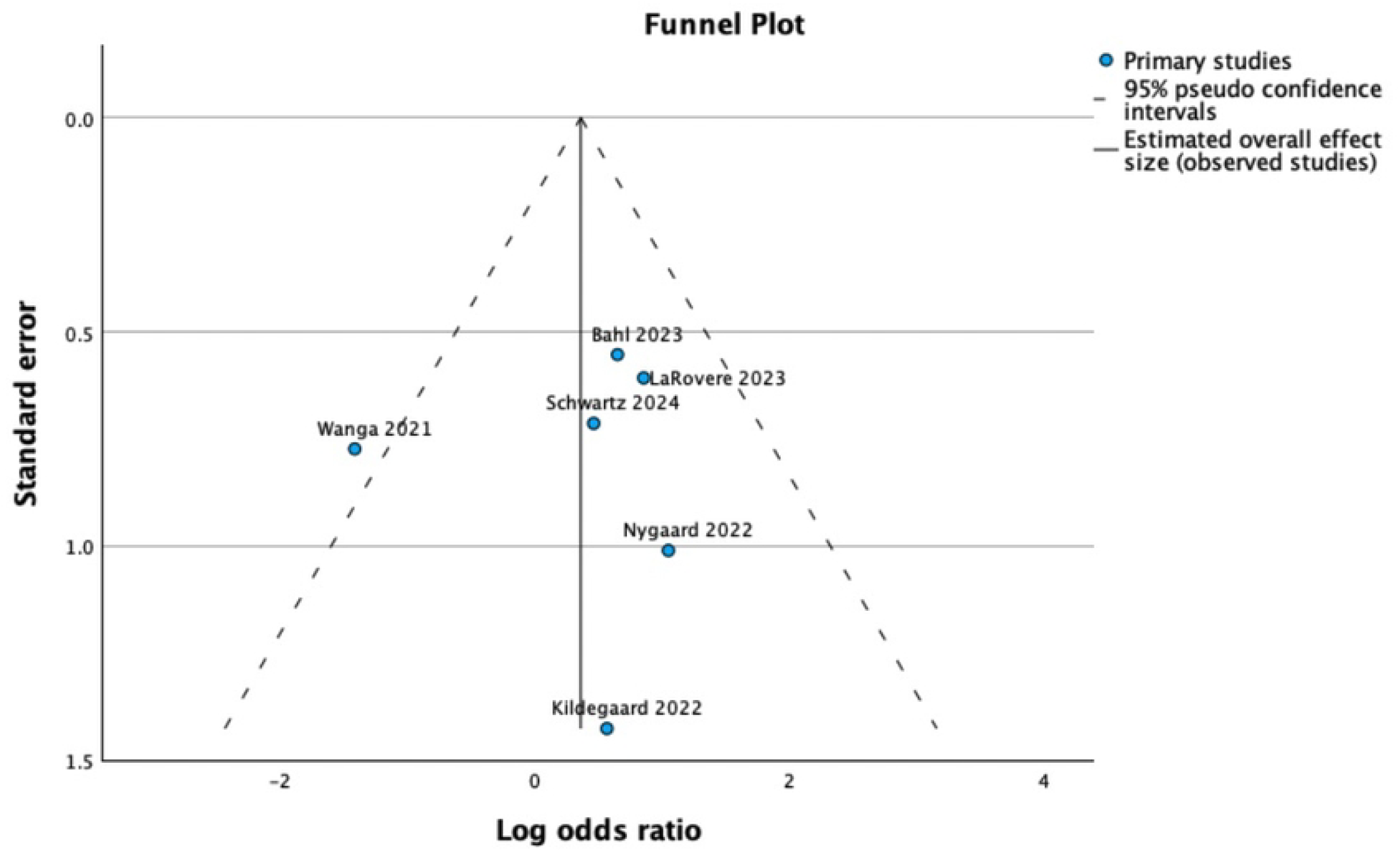
Funnel plot of the effect of two vaccinations on development of MIS-C, displaying an asymmetric graph implying potential publication bias.

The decision to conduct a sensitivity analysis excluding the Wanga et al. paper was informed by multiple methodological limitations identified during risk of bias assessment: (1) the shortest follow-up duration (one month) of all included studies, potentially insufficient to capture MIS-C onset; (2) the smallest vaccinated cohort (n=21), limiting precision; and (3) reliance exclusively on PCR or antigen testing without serological confirmation of prior infection, which may underestimate MIS-C incidence.

The analysis excluding Wanga et al. demonstrated a significant effect size, d = 0.71 [95% CI 0.07 to 1.35; p=0.03]. Heterogeneity was eliminated (I^2^ = 0%), and the funnel plot showed no evidence of publication bias (Figure 4).

**Figure 4a.**
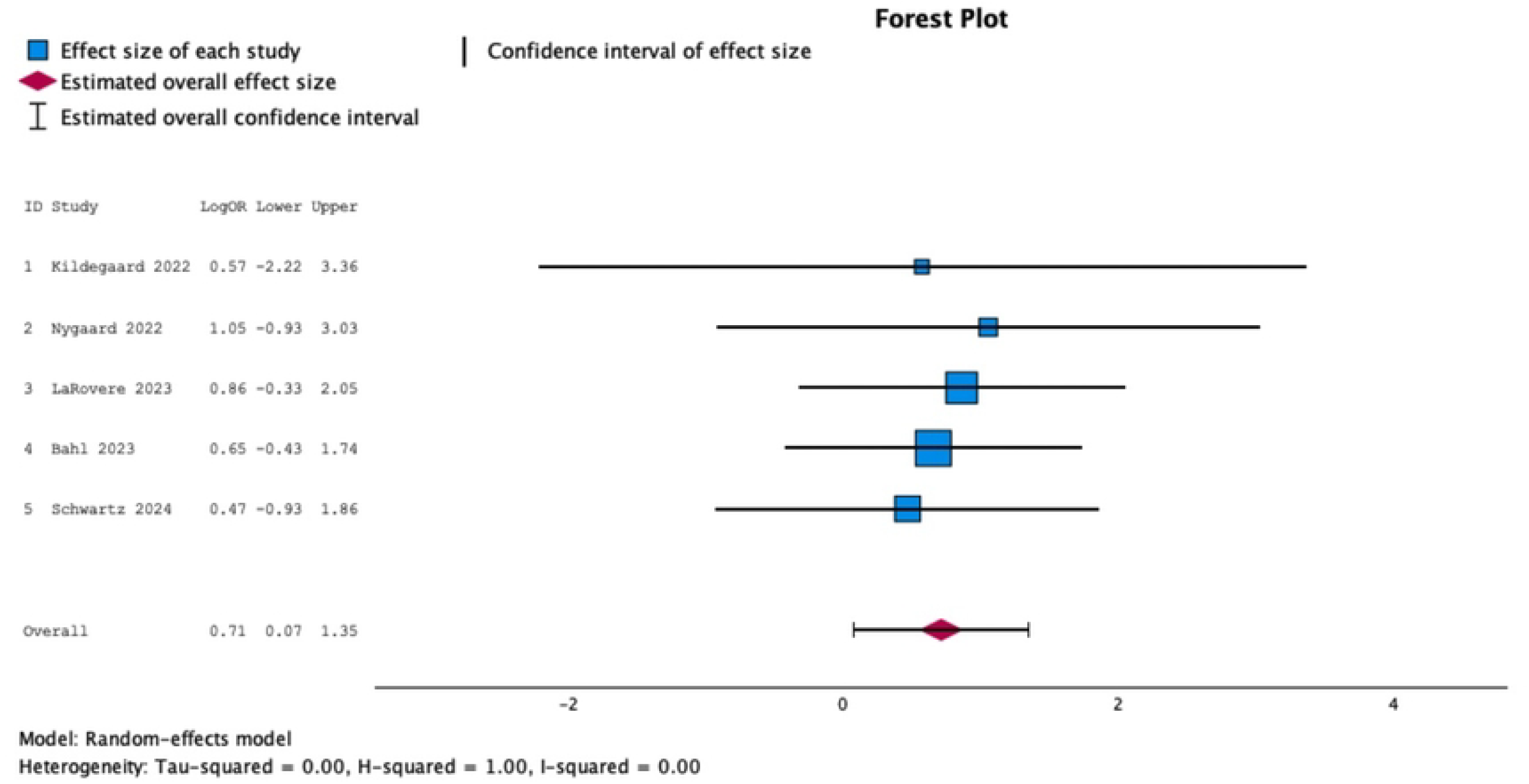
Forest plot of the effect of two vaccinations on development of MIS-C excluding Wanga et al, showcasing effect size of 0.77 (0.06∼1.49)

**Figure 4b.**
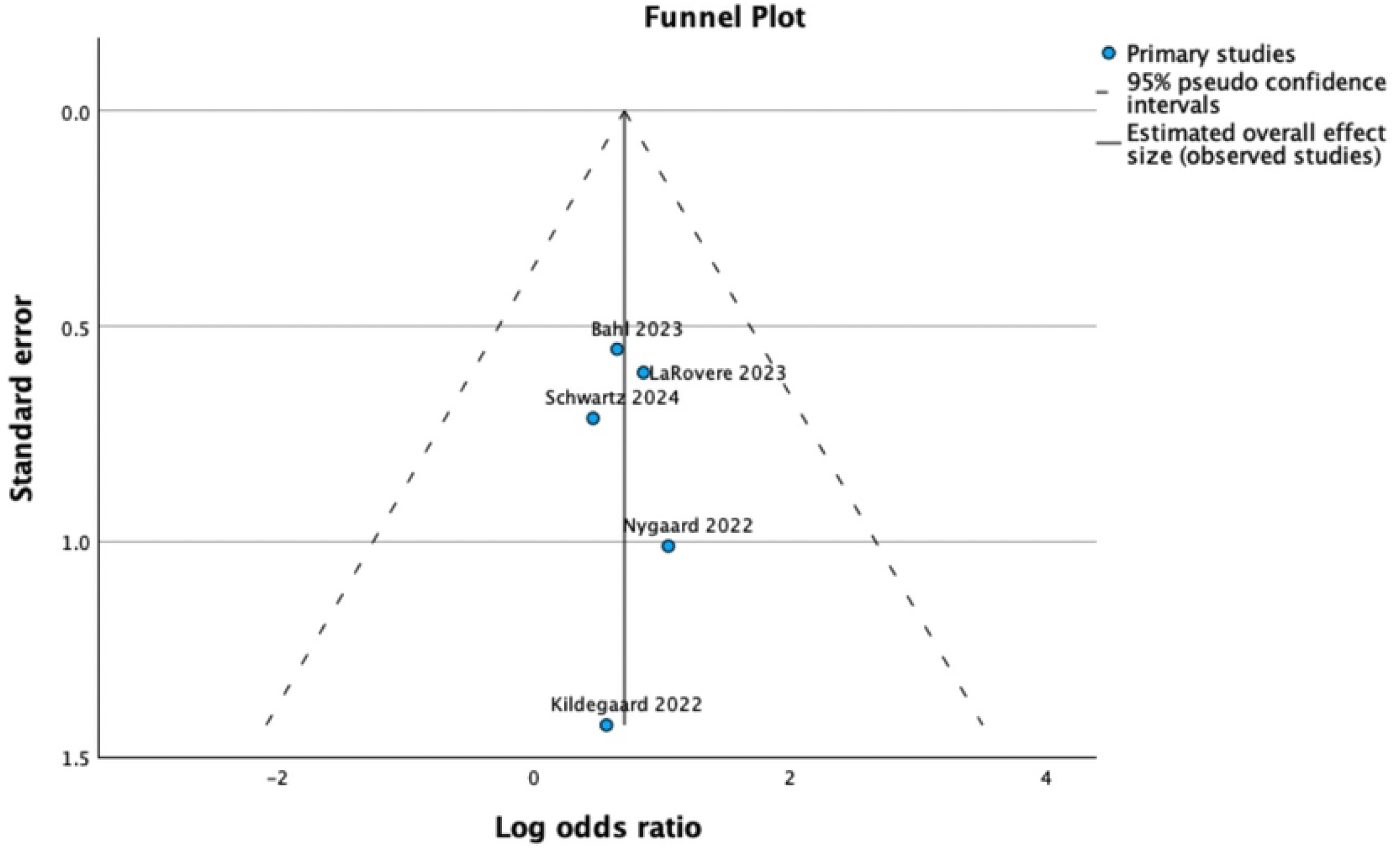
Funnel plot and forest plot of the effect of two vaccinations on development of MIS-C excluding Wanga et al., displaying a symmetric graph implying low publication bias.

### Assessment of Risk of Bias

Result of risk of bias assessment is represented in table 2.

## Discussion

The analysis of at least one dose of vaccination found a log odds ratio = 0.39 [95% CI - 0.17 to 0.95; p=0.17], with a heterogeneity I^2^ = 0% and a symmetric funnel plot. The result indicates a trend towards a protective effect that did not achieve statistical significance. The bias analysis is indicative of a non-biased study selection.

The initial analysis of two or more doses of vaccination versus less than two doses of vaccination found heterogeneity variance (tau squared) of 0.23 and I^2^ of 28%, indicating moderate heterogeneity effect beyond expected variability attributable to measurement error. Analysis also showed an asymmetric funnel plot, indicating potential selection bias. While the funnel plot suggested Wanga et al. study might be an outlier, the rationale for exclusion was primarily based on pre-identified methodological concerns. Specifically, Wanga et al. differed from the other studies in several aspects. First, the study was conducted 2-3 months after vaccination was approved for children of 12-15 years of age, contributing to a potentially skewed population base. Second, this paper included the smallest number of vaccinated children, with 21 out of 915 patients receiving at least one dose of vaccination. Third, this study has the shortest study duration of any paper included for analysis. Finally, this study used a positive PCR test or rapid antigen test only to identify MIS-C patients, which may have underestimated the MIS-C subset due to the lack of immunoglobulin serology results.

The sensitivity analysis excluding Wanga et al. showed a statistically significant protective effect of two doses of vaccination against MIS-C in breakthrough infections (d = 0.71 [95% CI 0.07 to 1.35; p=0.03]). Heterogeneity was eliminated (I^2^ = 0%), and the funnel plot demonstrated no evidence of publication bias. Our findings are consistent with prior observational studies. For example, Ouldali et al. *(37)* analyzed MIS-C incidence in vaccinated versus unvaccinated children and reported a substantially lower rate of MIS-C among vaccinated children, despite rare cases occurring after vaccination. This supports the interpretation that vaccination reduces MIS-C risk overall, even though breakthrough cases can still occur.

The result of this systematic review and meta-analysis indicates that the relationship between vaccination status and the odds of MIS-C incidence is not statistically significant in patients who are partially vaccinated. In the patient population who are fully vaccinated, there is a statistically significant protective effect after excluding a potentially biased sample, indicating that vaccination is protective against the incidence of MIS-C.

### Limitations and Future Directions

This systematic review and meta-analysis was limited primarily due to the number of available papers that met inclusion criteria. While numerous papers discussing MIS-C have been published, many were conducted by analyzing characteristics of all MIS-C cases in a certain database, and did not compare them to a SARS-CoV-2 positive cohort. While the most common vaccine in the analyzed studies was the Pfizer BNT162b2 mRNA vaccine, only three of the six included papers focused exclusively on children who had been vaccinated only with the BNT162b2. Two other studies included patients who had received any type of vaccination against SARS-CoV-2. Because of this heterogeneity and incomplete reporting, a vaccine-type specific analysis was not feasible and not attempted. In addition, most countries allowed older children to receive vaccinations first, later approving doses for progressively younger children. For example, in the United States, children aged 16 or older were eligible for vaccination beginning December 11, 2020, while children aged 12-15 became eligible on May 10, 2021, aged 5-11 on October 29, 2021, and aged 6 months to 5 years on June 17, 2022 *(38 - 41)*. This means that this analysis primarily draws conclusions about older children. As more papers are published regarding MIS-C cases in younger children, more expansive analysis can be conducted. To address these limitations, the database search was rerun multiple times to ensure the most complete data available. A further limitation of the analysis was sample size. Despite all the studies used in this analysis having a minimum of 500 subjects, MIS-C remains a rare complication, making large-scale analysis challenging.

With more data available for COVID-19 and MIS-C, a future direction would be to analyze the incidence of MIS-C across different waves of COVID-19. While this paper showcases the benefit of vaccination in prevention of MIS-C incidence, another future direction would be to conduct a meta-analysis on risk-benefit analysis of vaccination against MIS-C, as both MIS-C and side effects. In addition, research directly comparing vaccine-type-specific effectiveness, including mRNA and inactivated vaccines, in younger children under 12 years of age is needed to inform immunization strategies. These directions may help optimize both prevention and treatment approaches for MIS-C.

## Data Availability

No datasets were generated or analysed during the current study. All relevant data from this study will be made available upon study completion.

## Acknowledgments

The authors would like to acknowledge Doreen Olvet, PhD for her assistance in statistical analysis.

## Disclaimers

N/A.

**Author Bio** (first author only, unless there are only 2 authors)

Katherine Carroll, MD is an emergency medicine resident at the University of Cincinnati. Her current interests include medical education and administration.

## Footnotes (if applicable)

N/A.

## References

1. Cheung EW, Zachariah P, Gorelik M, et al. Multisystem Inflammatory Syndrome Related to COVID-19 in Previously Healthy Children and Adolescents in New York City. JAMA. 2020;324(3):294–296. doi:10.1001/jama.2020.10374

2. Verdoni L, Mazza A, Gervasoni A, et al. An outbreak of severe Kawasaki-like disease at the Italian epicentre of the SARS-CoV-2 epidemic: an observational cohort study. Lancet (London, England). 2020;395(10239):1771–1778. doi:10.1016/S0140-6736(20)31103-X

3. Whittaker E, Bamford A, Kenny J, et al. Clinical Characteristics of 58 Children With a Pediatric Inflammatory Multisystem Syndrome Temporally Associated With SARS-CoV-2. JAMA. 2020;324(3):259–269. doi:10.1001/jama.2020.10369

4. Riphagen S, Gomez X, Gonzalez-Martinez C, Wilkinson N, Theocharis P. Hyperinflammatory shock in children during COVID-19 pandemic. Lancet (London, England). 2020;395(10237):1607–1608. doi:10.1016/S0140-6736(20)31094-1

5. Gkoutzourelas A, Bogdanos DP, Sakkas LI. Kawasaki Disease and COVID-19. Mediterr J Rheumatol. 2020;31(Suppl 2):268–274. doi:10.31138/mjr.31.3.268

6. Radia T, Williams N, Agrawal P, et al. Multi-system inflammatory syndrome in children & adolescents (MIS-C): A systematic review of clinical features and presentation. Paediatr Respir Rev. 2021;38:51–57. doi:10.1016/j.prrv.2020.08.001

7. Santos MO, Gonçalves LC, Silva PAN, et al. Multisystem inflammatory syndrome (MIS-C): a systematic review and meta-analysis of clinical characteristics, treatment, and outcomes. J Pediatr (Rio J). 2022;98(4):338–349. doi:10.1016/j.jped.2021.08.006

8. Kabeerdoss J, Pilania RK, Karkhele R, Kumar TS, Danda D, Singh S. Severe COVID-19, multisystem inflammatory syndrome in children, and Kawasaki disease: immunological mechanisms, clinical manifestations and management. Rheumatol Int. 2021;41(1):19–32. doi:10.1007/s00296-020-04749-4

9. Sacco K, Castagnoli R, Vakkilainen S, et al. Immunopathological signatures in multisystem inflammatory syndrome in children and pediatric COVID-19. Nat Med. 2022;28(5):1050–1062. doi:10.1038/s41591-022-01724-3

10. Bartsch YC, Wang C, Zohar T, et al. Humoral signatures of protective and pathological SARS-CoV-2 infection in children. Nat Med. 2021;27(3):454–462. doi:10.1038/s41591-021-01263-3

11. Diorio C, Henrickson SE, Vella LA, et al. Multisystem inflammatory syndrome in children and COVID-19 are distinct presentations of SARS-CoV-2. J Clin Invest. 2020;130(11):5967–5975. doi:10.1172/JCI140970

12. Carter MJ, Fish M, Jennings A, et al. Peripheral immunophenotypes in children with multisystem inflammatory syndrome associated with SARS-CoV-2 infection. Nat Med. 2020;26(11):1701–1707. doi:10.1038/s41591-020-1054-6

13. Vella LA, Giles JR, Baxter AE, et al. Deep immune profiling of MIS-C demonstrates marked but transient immune activation compared to adult and pediatric COVID-19. Sci Immunol. 2021;6(57). doi:10.1126/sciimmunol.abf7570

14. Diorio C, Shraim R, Vella LA, et al. Proteomic profiling of MIS-C patients indicates heterogeneity relating to interferon gamma dysregulation and vascular endothelial dysfunction. Nat Commun. 2021;12(1):7222. doi:10.1038/s41467-021-27544-6

15. Gurlevik SL, Ozsurekci Y, Sağ E, et al. The difference of the inflammatory milieu in MIS-C and severe COVID-19. Pediatr Res. 2022;92(6):1805–1814. doi:10.1038/s41390-022-02029-4

16. Mistry P, Barmania F, Mellet J, et al. SARS-CoV-2 Variants, Vaccines, and Host Immunity. Front Immunol. 2021;12:809244. doi:10.3389/fimmu.2021.809244

17. Jiang L, Tang K, Levin M, et al. COVID-19 and multisystem inflammatory syndrome in children and adolescents. Lancet Infect Dis. 2020;20(11):e276–e288. doi:10.1016/S1473-3099(20)30651-4

18. Rosenberg ES, Dorabawila V, Easton D, et al. Covid-19 Vaccine Effectiveness in New York State. N Engl J Med. 2022;386(2):116–127. doi:10.1056/NEJMoa2116063

19. Olson SM, Newhams MM, Halasa NB, et al. Effectiveness of BNT162b2 Vaccine against Critical Covid-19 in Adolescents. N Engl J Med. 2022;386(8):713–723. doi:10.1056/NEJMoa2117995

20. Zheng C, Shao W, Chen X, Zhang B, Wang G, Zhang W. Real-world effectiveness of COVID-19 vaccines: a literature review and meta-analysis. Int J Infect Dis IJID Off Publ Int Soc Infect Dis. 2022;114:252–260. doi:10.1016/j.ijid.2021.11.009

21. Lin DY, Xu Y, Gu Y, et al. Effects of COVID-19 Vaccination and Previous SARS-CoV-2 Infection on Omicron Infection and Severe Outcomes in Children Under 12 Years of Age in the USA: An Observational Cohort Study.hTe Lancet. Infectious Diseases. 2023;23(11):1257–1265. doi:10.1016/S1473-3099(23)00272-4.

22. Simmons AE, Amoako A, Grima AA, et al. Vaccine Effectiveness Against Hospitalization Among Adolescent and Pediatric SARS-CoV-2 Cases Between May 2021 and January 2022 in Ontario, Canada: A Retrospective Cohort Study. PloS One. 2023;18(3):e0283715. doi:10.1371/journal.pone.0283715.

23. Yan VKC, Cheng FWT, Chui CSL, et al. Effectiveness of BNT162b2 and CoronaVac Vaccines in Preventing SARS-CoV-2 Omicron Infections, Hospitalizations, and Severe Complications in the Pediatric Population in Hong Kong: A Case-Control Study. Emerging Microbes & Infections. 2023;12(1):2185455. doi:10.1080/22221751.2023.2185455.

24. Zhang YF, Xia CY, Yang Q, et al. The Protective Effects of Pediatric Vaccination on Multisystem Inflammatory Syndrome in Children Stratified by Vaccine Status, Types and Virus Variants. International Immunopharmacology. 2023;125 (Pt A):111105. doi:10.1016/j.intimp.2023.111105.

25. Watanabe A, Kani R, Iwagami M, et al. Assessment of Efficacy and Safety of mRNA COVID-19 Vaccines in Children Aged 5 to 11 Years: A Systematic Review and Meta-analysis. JAMA Pediatrics. 2023;177(4):384–394. doi:10.1001/jamapediatrics.2022.6243.

26. Piechotta V, Siemens W, Thielemann I, et al. Safety and Effectiveness of Vaccines Against COVID-19 in Children Aged 5-11 Years: A Systematic Review and Meta-Analysis. The Lancet. Child & Adolescent Health. 2023;7(6):379–391. doi:10.1016/S2352-4642(23)00078-0.

27. Levy N, Koppel JH, Kaplan O, et al. Severity and Incidence of Multisystem Inflammatory Syndrome in Children During 3 SARS-CoV-2 Pandemic Waves in Israel. JAMA. 2022;327(24):2452–2454. doi:10.1001/jama.2022.8025

28. Miller AD, Yousaf AR, Bornstein E, et al. Multisystem Inflammatory Syndrome in Children During Severe Acute Respiratory Syndrome Coronavirus 2 (SARS-CoV-2) Delta and Omicron Variant Circulation—United States, July 2021–January 2022 . Clin Infect Dis. 2022;75(Supplement_2):S303–S307. doi:10.1093/cid/ciac471

29. Miller AD, Zambrano LD, Yousaf AR, et al. Multisystem Inflammatory Syndrome in Children—United States, February 2020–July 2021. Clin Infect Dis. 2022;75(1):e1165–e1175. doi:10.1093/cid/ciab1007

30. Kildegaard H, Lund LC, Højlund M, Stensballe LG, Potteg\r ard A. Risk of adverse events after covid-19 in Danish children and adolescents and effectiveness of BNT162b2 in adolescents: cohort study. BMJ. 2022;377. doi:10.1136/bmj-2021-068898

31. Nygaard U, Holm M, Hartling UB, et al. Incidence and clinical phenotype of multisystem inflammatory syndrome in children after infection with the SARS-CoV-2 delta variant by vaccination status: a Danish nationwide prospective cohort study. Lancet Child Adolesc Heal. 2022;6(7):459–465. doi:10.1016/S2352-4642(22)00100-6

32. LaRovere KL, Poussaint TY, Young CC, et al. Changes in Distribution of Severe Neurologic Involvement in US Pediatric Inpatients With COVID-19 or Multisystem Inflammatory Syndrome in Children in 2021 vs 2020. JAMA Neurol. 2023;80(1):91–98. doi:10.1001/jamaneurol.2022.3881

33. Bahl A, Mielke N, Johnson S, Desai A, Qu L. Severe COVID-19 outcomes in pediatrics: an observational cohort analysis comparing Alpha, Delta, and Omicron variants. Lancet Reg Heal - Am. 2023;18:100405. doi:10.1016/J.LANA.2022.100405

34. Wanga V, Gerdes ME, Shi DS, et al. Characteristics and Clinical Outcomes of Children and Adolescents Aged <18 Years Hospitalized with COVID-19 - Six Hospitals, United States, July-August 2021. MMWR Morb Mortal Wkly Rep. 2021;70(5152):1766–1772. doi:10.15585/mmwr.mm705152a3

35. Schwartz N, Ratzon R, Hazan I, et al. Multisystemic inflammatory syndrome in children and the BNT162b2 vaccine: a nationwide cohort study. Eur J Pediatr. Aug 2024;183(8):3319–3326. doi:10.1007/s00431-024-05586-4

36. Zambrano LD, Newhams MM, Olson SM, et al. BNT162b2 mRNA Vaccination Against Coronavirus Disease 2019 is Associated With a Decreased Likelihood of Multisystem Inflammatory Syndrome in Children Aged 5-18 Years-United States, July 2021 - April 2022. Clin Infect Dis. Feb 08 2023;76(3):e90–e100. doi:10.1093/cid/ciac637

37. Ouldali N, Bagheri H, Salvo F, et al. Hyper inflammatory syndrome following COVID-19 mRNA vaccine in children: A national post-authorization pharmacovigilance study. Lancet Reg Heal Eur. 2022;17:100393. doi:10.1016/j.lanepe.2022.100393

38. FDA. FDA Takes Key Action in Fight Against COVID-19 By Issuing Emergency Use Authorization for First COVID-19 Vaccine. https://www.fda.gov/news-events/press-announcements/fda-takes-key-action-fight-against-covid-19-issuing-emergency-use-authorization-first-covid-19. Published 2020. Accessed May 2, 2023.

39. FDA. Coronavirus (COVID-19) Update: FDA Authorizes Pfizer-BioNTech COVID-19 Vaccine for Emergency Use in Adolescents in Another Important Action in Fight Against Pandemic.

40. FDA. FDA Authorizes Pfizer-BioNTech COVID-19 Vaccine for Emergency Use in Children 5 through 11 Years of Age. https://www.fda.gov/news-events/press-announcements/fda-authorizes-pfizer-biontech-covid-19-vaccine-emergency-use-children-5-through-11-years-age. Published 2021. Accessed May 2, 2023.

41. FDA. Coronavirus (COVID-19) Update: FDA Authorizes Moderna and Pfizer-BioNTech COVID-19 Vaccines for Children Down to 6 Months of Age. https://www.fda.gov/news-events/press-announcements/coronavirus-covid-19-update-fda-authorizes-moderna-and-pfizer-biontech-covid-19-vaccines-children. Published 2022.

